# Understanding the Impact of Social Engagement Activities, Health Protocol Maintenance, and Social Interaction on Depression During Covid-19 Pandemic Among Older Americans

**DOI:** 10.1101/2023.02.04.23285479

**Authors:** Roungu Ahmmad, Paul A. Burns, Ashraful Alam, Jeannette Simino, Wondwosen Yimer, Fazlay Faruque

## Abstract

**Objectives:** Depression is a critical public health concern among older Americans. However, little is known about how older adults’ social engagement activities, health protocol maintenance, and social interaction (both physically and virtually) potentially contribute to their feelings of depression.

**Methods:** Data were collected from the Covid-19 supplement to the National Health and Aging Trend Study (NHATS) and core longitudinal follow-up study. A total of 3,181 Medicare-eligible older adults between June and December 2020 were examined how self-reported depression is related to social engagement activities, health protocols, social interaction with friends and family (F&F), and doctors using multiple logistic regression.

**Results:** This study reveals that the lack of social engagement activities, such as birthday parties and long-term care visits significantly contributes to older adults’ depression (OR: 1.34, 90% CI: 1.07-1.68, p=0.012, and OR: 1.28, 90% CI: 1.01-1.65, p=0.053 respectively). Subsequently, health protocols compliance with washing hands and wearing masks in public places were more likely 2.36 times and 3.44 times higher symptoms of depression for the older adults than those who were not maintaining those protocols (OR: 2.36, 90% CI: 1.24-4.57, p=0.009, OR: 3.44, 90% CI: 1.97-6.17, p<0.001 respectively). Furthermore, the lack of virtual social interaction via phone and text message with F&F is significantly related to depression whereas email or video call are not significantly related to depression for older adults. During Covid-19 pandemic, in-person visits with doctors significantly reduced patients’ depression on the other hand email communication significantly increased. However other virtual interactions with doctors did not significantly associate with patients’ depression.

**Conclusion:** The lack of social engagement, maintaining health protocols, and lacking virtual interactions over the phone significantly increase depression symptoms for older adults during the Covid-19 pandemic. Therefore, it would be beneficial to take initiative to engage older adults in a variety of social activities to make them feel more connected to their community. The older population should be contacted by phone during the Covid-19 pandemic with encouraging messages and informed of the importance of maintaining health protocols.

## Introduction

The Covid-19 virus poses a significant threat to physical and mental health among all ages, especially older people (Pfefferbaum and North 2020; Knudsen and Chokshi, 2020). Evidence has shown that infectious disease outbreaks are associated with immediate mental and emotional harm’ some of which abated over time, while others became persistent or chronic concerns, such as depression, anxiety, and so on disorders (Brooks et al., 2020; Rubin and Wessely, 2020). Worldwide, depression disorders have become more prevalent among all age groups for a variety of reasons (Firth et al., 2017; Kobbeltved et al., 2005; Carstensen et al., 2000; Grist and Cavanagh, 2013; GarcÍa-Lizana and Muñoz-Mayorga, 2010; Bruine de Bruin and Carman, 2018, ; Bruine de Bruin and Bennett, 2020).

The Centers for Disease Control (CDC) has reported that older adults are more likely to be infected with Covid-19 (Cleveland and Krista, 2021). Psychiatric symptoms and higher morbidities associated with Covid-19 and found that Covid-19 affected populations have had higher psychiatric symptoms, such as stress, exhaustion, anxiety, irritability, insomnia, etc. (Brooks et al., 2020, Panda et al., 2021). During the pandemic, anxiety and depression levels were higher than before the lockdown, especially among women, according to a Danish study (Panda et al., 2021). Additionally, another study found moderate-to-severe levels of subjective stress, anxiety, and depression occurred one month after lockdown (Robbins et al., 2021; Ustun, 2021). Also, a systematic review and meta-analysis conducted on l65 studies, primarily from China, relating to the mental health consequences of Covid-19 on the public and health professionals. This study found that Covid-19 had a negative effect on mental health and had a higher rate of morbidity and mortality for all aged populations (Holmes et al., 2020; Lai et al., 2020; Taquet et al., 2021).

Depression is now the second leading cause of disability worldwide (Murray et al.,2010). Several sociodemographic factors influence depression differences, including age, sex, marital status, race, and living circumstances (Bowung et al., 1991). However, due to Covid-19, state and federal restrictions on public gatherings and the maintenance of health protocols have significantly increased mental distress for all aged and race population (Lai et al., 2020; Verma and Mishra, 2020). Studies shown the majority of older adults are mostly dependent on their friends and family for various reasons and they are often in touch with them in person or virtually by phone call, text, e-mail, or video chat. However, with the advent of Covid-19, people spend less time interacting with older people (Freedman, Hu, & Kasper, 2021; Ankuda, Kotwal, Reckrey, Harrison, & Ornstein, 2022). Given that older adults were restricted to engage in social activities, strictly advised to maintain health compliances, and experienced limited physical social interaction. The purpose of this study was to examine how these factors could affect the mental health of older people, particularly depression, by shedding more light on their emotional state.

### Older adults and Covid-19

It is more likely that older people will suffer from serious illnesses and die, so they are likely to feel more fear and worry about dying as a result of Covid-19 (Verdery et al., 2021; Zaveri et al., 2020; Klinger et al., 2021; Dowd et al., 2020). The prevalence of Covid-19 has been found to be over 10% among those who are 80 years and older (Asahi et al., 2021; Verdery et al., 2021). There is evidence that the duration of quarantine that ranges from 2 to 66 days has a significant impact on depression and anxiety in older adults (Hawryluck et al., 2004). During the Covid-19 pandemic, quarantine times and lack of communication were even longer than usual. Mental disorders may be more prevalent among the older population. Therefore, there is a need to better understand the relationship between Covid-19 and depression among older individuals.

### Limiting social engagement and adverse emotional effects

Engagement is conceptualized as a multifaceted notion which lies in cognitive, affective, and behavioral dimensions (Johnston, 2018). The cognitive aspect of engagement specifies people’s positive and negative emotional state of anger, fear, enjoyment, support and belonging. In the context of social level engagement, it links to interaction, connection, involvement, and participation, among many other dimensions (Johnston, 2018). Community members could participate in an activity and interact with other members. People particularly the older adults’ demands for engagement in different social activities are perceived differently. Previous studies also unveiled empirical evidence on the relationship between older adults’ social engagement and their health consequences (Herzog, Ofstedal, & Wheeler, 2002). In an effort to control the infection rate of coronavirus, various restrictions have been imposed on individuals including social engagement activities, such as public gatherings, attendance in family obligating (e.g. birthday parties, weddings, or funerals, and visiting friends and relatives) were severely disrupted. Although the need for controlling the spread of Covid-19 is critical, the mitigation policies have created significant disruptions in daily life, such as spending less time out in public and less time socializing. The impact of these disruptions was acute for older adults, negatively impacting their emotional and psychological health and well-being (Barber and Kim, 2020; Courtemanche et al., 2021). Social disengagement might lead older adults to feel lonely. One study reported that around 19 percent of the participants stated the feeling of loneliness on more days during the pandemic (Barber and Kim, 2020). In specific, the decreasing of physical involvement and contact on the virtual platform including video calls with F&F, online engagement in classes, clubs, and other forms of activities were strongly related to the higher level of loneliness (Choi, Hammaker, DiNitto, & Marti, 2022). And loneliness among older people is found to be a cause for increased risk of poor health, limiting physical abilities such as performing daily living activities without help, and multiple diagnoses including depression, and hypertension, among many others (Jessen, Pallesen, Kriegbaum, & Kristiansen, 2018).

### Health Protocol Maintenance

Protective health-care behaviors are essential in order to control the risks of the spread of Covid-19. Some studies demonstrated that health protocol maintenance, such as physical distancing and musk-wearing behaviors contributed to reducing the Covid-19 spread (Howard et al., 2021). Since mortality risk among older adults was reported to be high, following risk mitigation behaviors was very important. Accordingly, people were strictly prescribed to follow health guidelines, including washing hands, maintaining a certain distance from other people, and wearing masks in public places. The rural older adults were less likely to maintain health-related protocols, such as keeping a six-feet distance, limiting gathering, avoiding bars or restaurants, avoiding touching faces, and avoiding in-person contact with outside people (Probst, Crouch, & Eberth, 2021). Given that older people are unlikely to maintain the health-related guideline, little research has addressed the issue by exploring its relation to their emotional outcomes, such as stress, depression, insomnia, and anger.

### Physical and Virtual Social Interaction

During the pandemic, there was a significant change in both the pattern of physical and virtual interactions. One study demonstrates that physical contact among older adults decreased substantially whereas virtual communication over telephone and electronic modes increased considerably during Covid-19 (Freedman, Hu, & Kasper, 2021). People were discouraged from engaging in in-person communication and instead encouraged to adopt virtual interactions through phone calls, text or e-mail messages, and video-calling. Research also shows that homebound older adults felt limited social contact due to Covid-19 (Ankuda et al., 2022). It also reported that older people were mostly contacted through email, text, and social media during the pandemic. Importantly, one study revealed that older adults experienced an increased rate of loneliness and depression (Ankuda et al., 2022) during the pandemic situation because of a lack of communication activities. However, little is known about how older adults’ limited physical social interaction and dependency on virtual social interaction respectively affect their mental health during Covid-19 pandemic.

### Proposing Research Questions

There were unavoidable barriers to daily living posed by the Covid-19 pandemic, including limited access to social activities, health-protocols maintenance, and some restrictions on physical social interaction. Given this, it is assumed that the policies of curtailing a person’s engagement in social engagement activities, imposing health-related guidelines, and restraining physical and virtual interaction could contribute to producing feelings of depression among older adults. Against this backdrop, this study addresses the following leading research question: How do social engagement activities, health protocol maintenance, and physical and virtual social interaction affect depression in older adults?

## Methods

### Research design, participants, and study framework

The present investigation is a cross-sectional study comprised of community-dwelling participants in the first wave of the NHATS release 10 final data and Covid-19 supplementary dataset. The NHATS is the successor to the National Long Term Care Survey and is a panel study of Medicare beneficiaries aged 65 and older population in the United States (Davydow et al., 2014). The NHATS used a stratified three-stage sample design with sampling based on U.S. county and residential zip code as well as age (Davydow et al., 2014) and the final sample included participants from every state except Alaska, Hawaii and Puerto Rico (Davydow et al., 2014). Participants and/or their proxies were interviewed in-person in 2020 through June to December, with annual reinterviews planned. The NHATS protocol was approved by the Johns Hopkins University Institutional Review Board, and all participants provided informed consent.

### Outcome variable

We used the four-item NHATS depression scale in this study to determine self-reported depression levels. The purpose of the NHATS is to develop standardized, accurate, valid, and reliable measures of patient-reported outcomes in the areas of physical, mental, and social health (Murray et al., 2010). In order to achieve greater precision and reduce respondent burden, the depression scale was developed based on item response theory (Murray et al., 2010). This scale has been cross-validated with other depression instruments, such as the Center for Epidemiological Studies Depression Scale (CES-D), the Beck Depression Inventory (BDI-II), and the Patient Health Questionnaire (PHQ-9) (Dadfar et al., 2021; Jiang et al., 2019; Maroufizadeh et al., 2019). A four-item depression scale asked participants how frequently they had felt hopeless, worthless, helpless, or depressed in the past six months. Those items were scored on a Likert scale ranging from 1 to 4, which corresponded to the response options “Not at all,” “Mild,” “Moderate” and “Severe”. Based on the non-normal distribution of data, raw scores were collapsed into two groups: “Not at all” means “No depression” and all other together is “Depression”. This was in line with the purpose of the depression scale, which was to grade severity rather than simply provide a clinical cut-off point. This classification was valid based on both the distribution of data and the clinical cut-off for depression recommended by the American Psychiatric Association (Dbouk et al., 2021; Choi et al., 2021).

### Covariates

For our analysis, the factors were clustered into four groups: 1) socio-demographic factors (age, sex, race, marital status), 2) social engagement activities (missed activities, such as birthday party, funeral, leisure, long trips, wedding ceremony), 3) health protocol maintenance (e.g., hand washing, maintained six feet distance in public places and mask wearing in public gatherings), and 4) physical and virtual social interaction with F&F and doctors (e.g., in-person meeting, phone call, e-mail/text message and video call).

### Statistical analyses

This study assessed multivariable associations and some hierarchical analyses of adults’ self-reported depression with their supportive networks after controlling for key demographic factors (i.e., age, sex, race and marital status). All participants with complete data on both the independent and dependent variables were included in all analyses. To describe our samples, we computed percentages of all the covariates and the dependent variables (depression). Chi-square tests were conducted to determine bivariable associations between independents and outcomes variables. We examined univariate and multivariate associations between depression and each predictor and to incorporate all covariates into our multivariable models. Finally, we considered some hierarchical analyses by adjusting demographic factors for a better understanding of the association of predictors with depression. Our statistical analyses were conducted with *R* programming language and *Stata* 15.0.

## Results

### Descriptive Analysis

Approximately 70% of the participants (n = 2,228) experienced depression (mild, moderate, or severe) in this study. Most respondents (58%) were female, where 76% were white, 24% were black respondents. Among the respondents, 96% were married, separated, or divorced, and only 4% were never married or single. Around 46% of the respondents were aged 60 to 80 years and 41% were age between 80 to 90 years. According to the results, about half of those surveyed missed a birthday party, funerals and long-term care visits. More specifically, around 51% (n = 1492,) people missed a birthday party, 41% (n = 1,154) missed a funeral, 36% (n = 1,023) missed long-term care visits, where around 74% of them suffered from depression (Table 1).

**Table 1.**
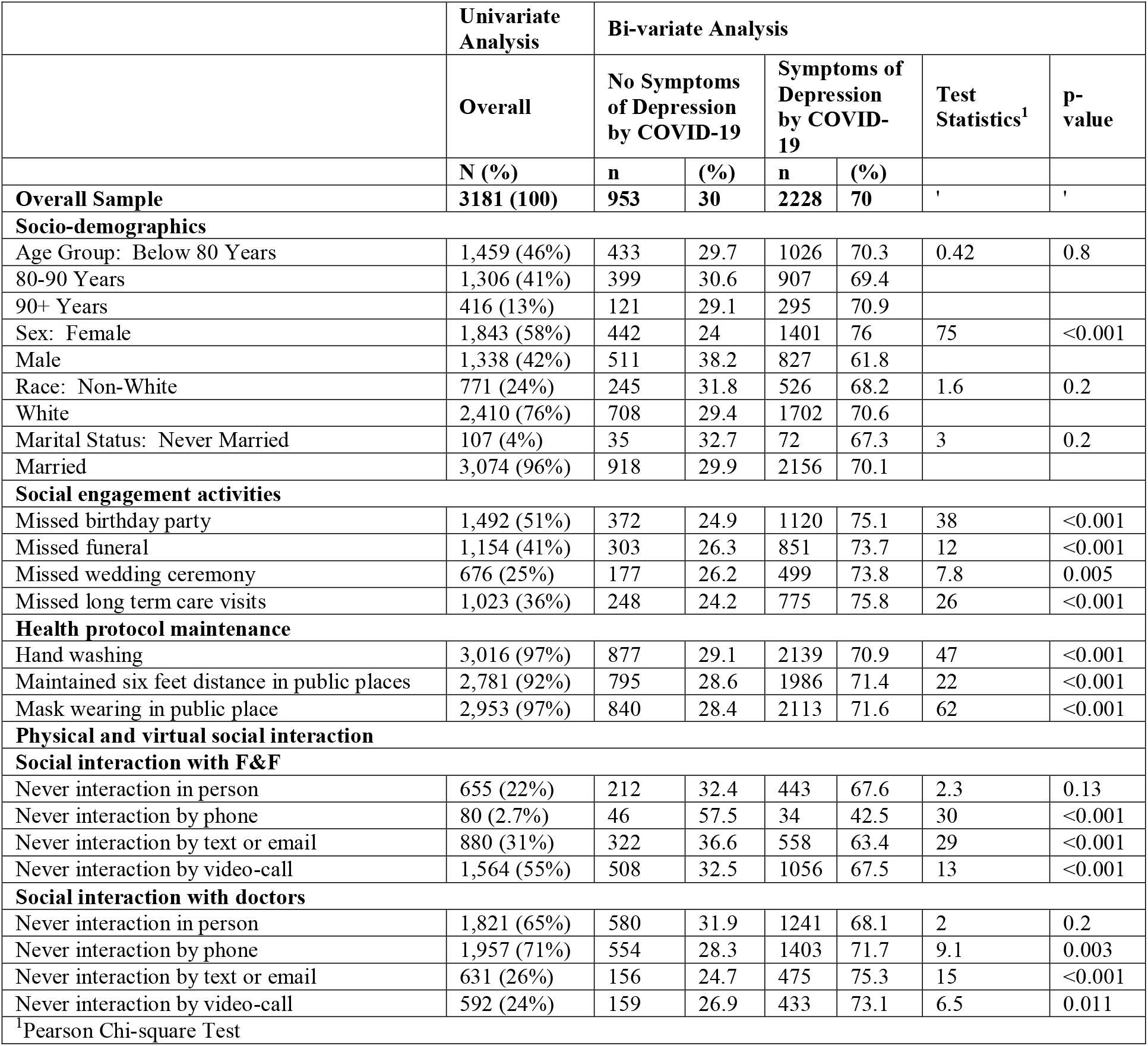
Univariate and bivariable analyses of the relations of social engagement activities, health protocols maintenance and physical and virtual social interaction of older adults with depression during the pandemic of COVID-19 (*N* = 3181).

Furthermore, this study found older female adults (n = 1401, 64%) were significantly more likely to report depression than their male counterparts (n = 827, 37%). Additionally, 90% of participants reported being able to maintain routine Covid-19 health protocols; 97% for hand washing and mask wearing in public places and 92% maintenance of six feet distance in public places. In bivariate analysis, we found those who reported depression were less likely to maintain Covid-19 protocols compared to those who were depressed. Among the people who missed birthday party, 75.1% felt depressed compared with only 24.9% had no system of depression (p <0.001). Similarly, for missed funeral, wedding ceremony and long-term care visit significantly increased feelings of depression. Health care protocol (hand washing, six feet distance and mask wear) also contributed to increasing depression during covid-19 pandemic. The lack of in-person social interactions with F&F and doctors were not significantly associated with the feeling of depression. But the absence of virtual social interaction with F&F and doctor (e.g., interaction via text or email) was significantly associated with feeling of depression for the older adults.

To address the leading RQ, we conducted adjusted logistic regression to examine the association between the social engagement activities and symptoms of depression during Covid-19 pandemic. According to the adjusted model, missing a birthday party (OR: 1.34, 90% CI: 1.07-1.68, p = 0.012), and a long-term care visit (OR: 1.28, 90% CI: 1.00-1.65, p = 0.053) were significantly increased the symptom of depression. In addition, older adults who reported washing hands during Covid-19 were more than twice as likely to report depression (OR: 2.36, 90% CI: 1.24-4.57, p = 0.009) than those who were not practicing hand washing. Similarly, this study also found that the older adults who wore masks in public places experienced 3.4 times higher rate of depression (OR: 3.44, 90% CI: 1.97-6.17, p = 0.001) than those who did not wear masks in public places (Table 2). Moreover, absence of social interactions with F&F via the phone (OR: 1.88, 90% CI: 0.95-3.73, p = 0.069) was significantly increase the symptom of depression for older adults in the pandemic time, but in-person meeting, e-mail/text and video-calls did not significantly associate with depression. Interactions with physicians via in-person meeting significantly decreased depression among older adults (OR: 0.82, 90% CI: 0.66-1.02, p=0.075); on the other hand, absence of virtual interaction did not significantly link with the symptom of depression.

**Table 2:**
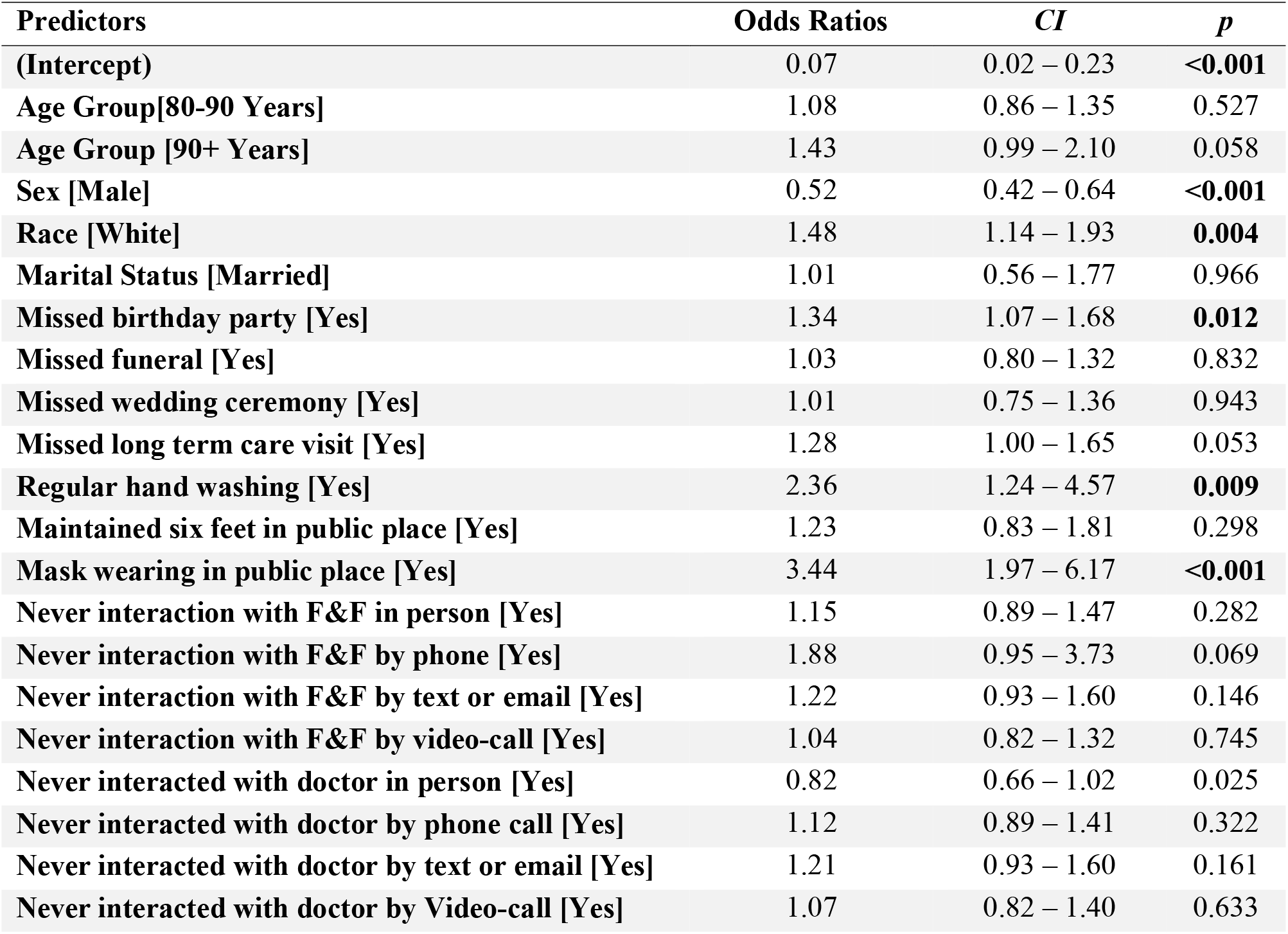
Adjusted logistic regression outputs of the relations of various factors of social engagement activities, health protocols maintenance and physical and virtual social interaction of older adults with depression during the pandemic of COVID-19 (*N* = 3181).

For better understanding the association between social engagement activities, health protocol, and social interaction with F&F and doctors, we fitted a multilevel hierarchical logistic regression model. We fitted four separate models for investigating the detail association between depression and predictors social engagement, health protocol, interaction with F&F and doctors. According to the hierarchical logistic regression in Figure 1, P1, missing birthday parties and long term care visit were significantly increased symptoms of depression (OR: 1.46, CI: 1.17-1.81, p <0.001). Additionally, people who regularly washed their hands because of COVID-19 had 2.68 times higher odds of being depressed than people who did not (OR: 2.68, 90% CI: 1.42-5.16, p = 0.003). A similar finding was found for wearing masks in public places (OR: 3.87, 90% CI: 2.24 – 6.88, p<0.001). The absence of social interaction with F&F by in-person and video call did not significantly contribute to the symptom of depression. However, lack of virtual social interaction over phone, email, or text message were significantly associated with depression (OR: 2.26, 90% CI: 1.10-4.45, P = 0.016, OR: 1.35, 90% CI: 1.04-1.74, P = 0.023). We also found that people who were able to communicate with doctors in person were less likely to be depressed compared to people who were unable to do so (OR: 0.82, 90% CI: 0.66-1.01, p=0.061). Never getting feedback from doctors via text messages or email significantly contribute feel of depression for the older adults (OR: 1.40, 95% CI: 1.09-1.82, p = 0.010). On the other hand, never having contact with phone call and video calls did not have a significant association with the symptom of depression for older adults.

**Figure 1:**
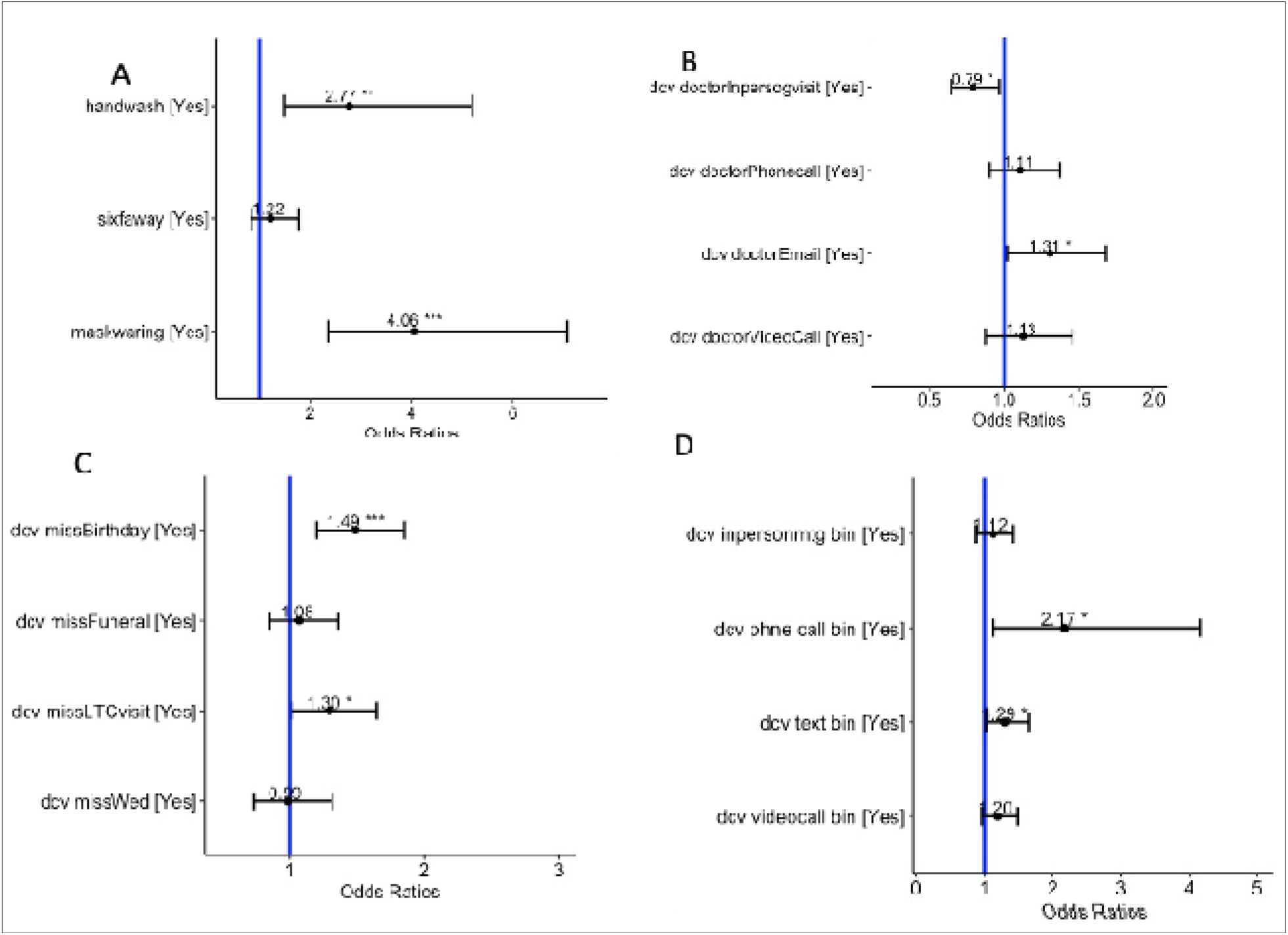
**Stratified or hierarchical logistic regression model by adjusting socio-demographic factors, P1: Social engagement activities, P2: Health protocol maintenance, P3: Physical and virtual social interaction with** F&F, **and P4: Physical and virtual social interaction with doctors.]**

### Discussion and Conclusion

This study investigated the relations of self-reported depression symptoms with the social engagement, health protocol and physical and virtual social interaction among older adults during the Covid-19 pandemic. Utilizing a nationally representative sample, this study found that depression is alarmingly common health outcomes among older adults. Nearly three quarters of older adults suffer from depression and the prevalence of substantial depression among community-dwelling American 65 years or older presented is consistence in previous study (Li C et al., 2007). This study also revealed that social engagement activities, health protocols maintenance and physical and virtual social interaction with F&F and doctors, play a vital role in explaining the depression of older adults caused by the Covid-19 pandemic. Missing activities of social engagement (e.g., birthday parties, long-term care visits), maintaining health related protocols (e.g., hand washing, mask wearing in public places) and the absence of social virtual interaction with F&F (e.g., via over phone) are strongly associated with symptom of depression among older adults.

Study shown that social gathering and interaction with F&F and discipline life relief mental stress and smooth life (Site et al., 2022; Sedig and Parsons, 2013). This study results showed that older adults who did not engage in social activities were more likely to report being depressed. During the pandemic, many older adults experience disruptions to their daily routines creating barriers to social and community engagement, which are key factors that have been shown to cause depression, particularly in the older population (Sim et al, 2021). Sleeping disturbance and changing daily activity significantly increased depression and anxiety during Covid-19 time among older adults (Rebecca et al., 2021). Consistent with previous study, results after adjusting demographic data, we found missing birthday parties and long-term care visits significantly increased symptoms of depression for the older adults.

In addition, this study contributed to expanding our understanding of maintaining health protocols during pandemics. The association between cognitive impairment and social determinants of health has been examined by other studies as well (Lim et al., 2018; Bressington et al.,2020; Torales et al., 2020; Rajkumar, 2020). In our study, we found that maintenance of health protocols like washing hands and wearing masks in public places was strongly associated with the symptom of depression. It is possible that the maintenance of related Covid-19 guidelines is prohibitive, resulting in added stress for older adults that can have detrimental effects on their mental health.

Importantly, the type and method of communication played a significant role in explaining depression among older adults during Covid-19. For example, older adults who interacted using virtual methods of communication (i.e. interaction with F&F via over phone call) were less likely to report depression. Consequently, the absence of phone calls and other contacts with F&F may increase feelings of depression. The case of in–person communication with doctors would be somewhat curtailed because of the possibilities of virus contamination. But the absence of in-person meeting with doctors seems to contribute to the depression of older adults. One possible reason might be that physical visit to the doctors is indispensable especially in the case of physical illness. Some other research indicated that electronic media communication now a day easy for older people attached with F&F via phone call, text or even video call, which makes them in touch with F&F (Brent et al., 2008; McLaughlin, 2011; Avenevoli et al., 2015). Our study found that never interaction with phone call was a significant factor for increasing the symptom of depression during Covid-19 time. However, in-person meeting and virtual social interactions via text message or video call were not significantly associated with the feeling of depression.

This study extended the literature on depression and older populations by unpacking the roles of some important factors, including social engagement activities, health related protocols, and physical and virtual social interactions. According to a nationally representative cohort of older adults aged 65 to 90+, depression is alarmingly prevalent among community-dwelling older adults during Covid-19. Even though this study used different ascertainment methods from previous studies (Lai et al., 2020; Ustun, 2021; Robbins et al., 2021; De Bruin, 2021; Harden et al., 2020), it found similar prevalence rates of probable depression among community-dwelling older Americans, which suggests reliability in our results and broadly consistent with previous studies. This study is an important contribution to literature as it suggests that some activities of social engagement, health protocols, and social interaction with family are necessary to consider for the mental health of the older population in the United States during the Covid-19 pandemic.

### Limitations and recommendations

There are several limitations in this study. First, this is a cross-sectional study and therefore we are unable to ascribe causality. Also, since only a small number of participants in NHATS completed the Covid-19 supplement, it may not fully reflect the U.S. population, therefore it may not be generalizable. Furthermore, the results should be interpreted with due caution as the questions regarding depression are not clearly stated to detect clinically relevant depression measurements because of the lack of clinical predictors in this study introduced. Instead of the above gaps, the findings of this study underlines the significance of social engagement of adult people in various activities for their mental wellbeing. Even though coronavirus put a bar in physical social engagement, initiative should be taken to arrange some social activities even virtually. Importantly, this research helps us comprehending why older adults unlikely want to maintain health protocols including wearing masks. Therefore, messages related to the health compliances would be conveyed across the country with persuasive ways. Additionally, as much as the older adults will be connected through increasing level of virtual social interactions, the depression would turn into happiness.

## Data Availability

NHATS website

https://www.nhats.org/

## Author Agreements

No author contradicted with this manuscript for publishing

## Acknowledgement

The authors acknowledge the NHATS for giving the opportunity to analysis the data.

